# A new diagnostic pipeline for recurrent diffuse podocytopathies post transplantation using glomerulus-on-a-chip technology

**DOI:** 10.64898/2026.11.27.26368810

**Authors:** Alice M Gage, William J Mason, Laura Perin, Stefano Da Sacco, Marina Loucaidou, Jonathan Briggs, Jennifer C Chandler, Karen L Price, David A Long, Alan D Salama

**Affiliations:** Developmental Biology and Cancer Research and Teaching Department, Great Ormond Street Institute of Child Health, Faculty of Population Health Sciences, University College London, London, UK; University College London Centre for Kidney and Bladder Health, Division of Medicine, Faculty of Medical Sciences, University College London, London, UK; Royal Free Hospital, University College London Medical School, Faculty of Medical Sciences, University College London, London, UK; GOFARR Laboratory for Organ Regenerative Research and Cell Therapeutics in Urology, Division of Urology, The Saban Research Institute, Children’s Hospital Los Angeles, Los Angeles, California, USA; Keck School of Medicine, University of Southern California, Los Angeles, California, USA; Hammersmith Hospital, London, UK

**Keywords:** antibody-associated kidney disease, focal segmental glomerular sclerosis, organ-on-a-chip technology, recurrent diffuse podocytopathy, therapeutic stratification pipeline, transplantation

## Abstract

**Introduction:** Recurrent diffuse podocytopathies in kidney transplants are a leading cause of graft loss with poorly understood pathophysiology and variable response to therapy. Recent work has focussed on anti-slit diaphragm antibodies, but reliable methodology for their detection has proven challenging. In addition, antibody-associated disease only accounts for disease mechanism in a proportion of patients. Clinicians need a reliable stratification pathway, firstly, to determine if a circulating factor indiscriminate of its nature is present, and secondly, to establish whether it is antibody-associated or otherwise, to facilitate a precision medicine approach to therapeutic management and transplantation.

**Methods:** This exploratory cohort study used primary glomerular cells within a modified Glomerulus-On-A-Chip (GOAC) model. To identify the presence of a pathogenic circulating factor, we exposed the GOAC to an inception cohort of healthy control (n=4), non-recurrent (n=4) and recurrent disease sera (n=14); assessing permeability by the addition of fluorescein isothiocyanate conjugate (FITC) albumin. To determine if recurrent disease was antibody-associated or otherwise, immunostaining was performed on sera-exposed GOACs and post-recurrence allograft biopsies. Subsequently, we validated our pipeline on six sera blinded to clinical outcome.

**Results:** GOAC exposure with all recurrent disease sera resulted in an increase in albumin permeability compared with healthy controls. The mean fold difference between recurrent and non-recurrent disease sera permeability was 3.42 (p <0.001). An anti-nephrin antibody-associated pattern was found by immunostaining for 9/13 (69%) recurrent disease sera and was consistent with individual clinical response to antibody-targeted therapy. ROC curve analysis showed excellent sensitivity and specificity of our pipeline (AUC 0.977, p <0.0001) which accurately predicted clinical outcomes for all six patients in the blinded validation cohort.

**Conclusions:** This functional GOAC pipeline accurately determines the presence of a pathogenic circulating factor and can define it as anti-nephrin-associated or otherwise to guide therapeutic strategy and optimise approaches to transplantation.

**Translational Statement:** Diffuse podocytopathies are conditions where damage to podocytes leads to urinary protein leak and reduced kidney function. Only 50% of patients respond to treatment, the rest develop CKD, some needing dialysis or transplantation. However, the disease returns in approximately 40% following transplantation due to circulating factor(s). Current tests for these are unreliable. Even though some are thought to be due to antibodies to the podocyte proteins, antibody-directed treatment only helps a minority recover. Our pipeline provides a translational strategy to detect the presence of a damaging circulating factor and determine whether it is an antibody to better inform patient care.

## Introduction

Recurrent diffuse podocytopathies (DP) are a leading cause of kidney allograft loss and remain one of the last disease related barriers to transplantation.^1,2^ The recurring phenotype indicates the involvement of a pathogenic circulating factor, but whether this is antibody-associated or otherwise is unknown in many cases.^3^ Incomplete understanding of the pathophysiology underpins the limited efficacy of current therapeutic strategies resulting in suboptimal patient outcomes.^1^

In native podocytopathies, there is an expanding body of work implicating slit diaphragm autoantibodies, most commonly anti-nephrin, but also anti-podocin and anti-KIRREL-1.^4–8^ The largest European study to date found 44% of adults with minimal change disease (MCD) and just 9% with focal segmental glomerulosclerosis (FSGS) had anti-nephrin antibodies leaving the majority with an unexplained mechanism of injury.^4^ In addition, reliable detection methods have remained challenging likely due a combination of factors including low and fluctuant antibody titres; heavily glycosylated recombinant slit-diaphragm antigens resulting in non-specific binding; and potential epitope spreading in severe disease.^9–11^

There is currently a paucity of evidence regarding anti-slit diaphragm antibodies in recurrent DPs with a significant range in reported prevalence. A single paediatric study demonstrated anti-nephrin antibodies in 14/14 patients whilst the only two studies in adult patients reported positive findings in 8% and 38% patients respectively.^12–14^ A more recent study, with a combined adult and paediatric population, found autoantibodies in 30% of recurrent disease cases.^5^ None of the recurrent FSGS patients were found to have an isolated anti-nephrin antibody, instead, anti-podocin or an anti-KIRREL 1 antibodies were detected, with or without additional anti-nephrin antibodies, with one patient demonstrating triple positivity.^5^ Anti-podocin and anti-KIRREL 1 positivity was associated with early disease recurrence and increased disease severity supporting the concept of epitope spreading in advanced recurrent FSGS and potentially implicating different antibody sensitivities to standard therapies.^5^

Therapeutic options for recurrent DPs are currently largely limited to plasma exchange (PEX) and B cell-targeting therapies (e.g. rituximab), the latter specifically addressing antibody-driven disease. Reported outcomes are variable: in the largest retrospective study to date, only one fifth achieved complete remission following combined PEX and rituximab therapy, with no apparent additional benefit in those receiving further doses of rituximab.^1^ A handful of case studies have reported success with off license use of daratumumab suggesting enhanced targeting of long-lived plasma cells.^15–19^ This leaves the majority without a defined pathological mechanism or an evidence-based strategy for treatment, frequently resulting in graft loss.^1^

To address this unmet need, we created a functional diagnostic pipeline to 1) identify patients at risk of recurrence due to a persistent circulating factor indiscriminate of its nature 2) describe the circulating factor as being anti-nephrin antibody associated or otherwise. This work allows us to identify patients at high risk of disease recurrence and to stratify them according to their likely response to B cell targeting therapy, informing individualised therapeutic decision making and maximising the likelihood of successful transplantation.

## Methodology

### Patient samples

Our inception cohort consisted of 22 adult participants: 14 patients with biopsy-proven recurrent DPs (FSGS (n=10), MCD (n=4)), 4 patients with non-recurrent DPs (n=4 FSGS) and 4 healthy controls (HC). Our validation cohort contained 6 adult participants, all with biopsy-proven recurrent FSGS. For all patients, sera were obtained at the point of crossmatch and in addition for 4 patients pre- and post-treatment (PEX and rituximab) samples were obtained.

Samples were collected in EDTA tubes and centrifuged at 2000 rpm for 10 minutes. Sera were transferred to 1.5 ml tubes and stored at –80°C. On the first thaw, samples were aliquoted, and each aliquot was freeze-thawed a maximum of 3 times. Post-recurrence biopsies were available for 8/14 patients. All samples (Table 1) were obtained with ethical approval (05/Q0508/6 Royal Free Hospital Research Ethics Committee (REC). Response to treatment was defined as uPCR <100 mg/mmol with stable creatine over 3 months; partial response as uPCR >100 < 300 mg/mmol with stable creatinine over 3 months; no response as uPCR >300 mg/mmol, with ongoing decline in function or return to renal replacement therapy due to recurrent disease.

**Table 1:** Patient demographics and clinical data. Table defining the sex, age, ethnicity, native diagnosis, pre-emptive treatment, treatment, and response to treatment. NR1-4 – inception cohort non-recurrent disease patients 1-4; R-1 – R-14 – inception cohort recurrent disease patients 1-14; V1-V6 – validation cohort patients 1-6. Response to treatment defined as uPCR <100 mg/mmol with stable creatine over 3 months, partial response uPCR >100 < 300 mg/mmol with stable creatine over 3 months, no response uPCR >300 mg/mmol, with ongoing decline in function or return to renal replacement therapy due to recurrent disease. (FSGS – focal segmental glomerular sclerosis, MCD – minimal change disease, PEX – plasma exchange, RTX – Rituximab, RASi – renal angiotensin system inhibition, Rx - treatment).

| Patient no. | Sex | Age | Ethnicity | Native diagnosis | Pre-emptive Rx? | Treatment | Response to Rx? |
|---|---|---|---|---|---|---|---|
| NR1 | M | 70’s | White | Primary FSGS | No | N/A | N/A |
| NR2 | M | 40’s | White | Primary FSGS | No | N/A | N/A |
| NR3 | F | 50’s | White | Primary FSGS | No | N/A | N/A |
| NR4 | F | 40’s | Mixed Race | Primary FSGS | No | N/A | N/A |
| R-1 | M | 40’s | White | Primary FSGS | No | PEX, RTX, abatacept | Partial |
| R-2 | M | 50’s | White | MCD | No | PEX, RTX | No |
| R-3 | M | 40’s | Asian | Primary FSGS | No | PEX, RTX | Yes |
| R-4 | F | 60’s | White | MCD | No | PEX, RTX | Yes |
| R-5 | M | 40’s | Mixed Race | Primary FSGS | No | PEX | Partial |
| R-6 | M | 40’s | White | Primary FSGS | No | PEX, RTX | Yes |
| R-7 | M | 30’s | Asian | Primary FSGS | No | No | Yes |
| R-8 | F | 60’s | White | Primary FSGS | No | PLEX | Yes |
| R-9 | M | 70’s | White | MCD | Yes - indomethacin | Nil - nephrectomy | N/A |
| R-10 | M | 50’s | Black | Unknown | No | Increased RASi | Partial |
| R-11 | F | 50.s | Black | Unknown | No | PEX, RTX | Partial |
| R-12 | F | 30’s | Black | Primary FSGS | Yes - x1 PEX | PEX, RTX | Yes |
| R-13 | M | 30’s | Black | Primary FSGS | Yes - x1 PEX + 2g RTX | Increased RASi | Yes |
| R-14 | F | 40’s | White | MCD | No | PEX, RTX | Partial |
| V-1 | F | 40’s | White | Medullary Sponge Kidney | No | PEX, RTX | No – PEX dependent |
| V-2 | F | 20’s | White | Primary FSGS | Yes – x1 PEX | PEX, Obinutuzumab | Partial – PEX dependent |
| V-3 | M | 40’s | Asian | MCD | No | PEX, RTX | Yes |
| V-4 | M | 50’s | Black | Primary FSGS | No | PEX, RTX | No |
| V-5 | M | 60’s | Black | Primary FSGS | No | PEX, RTX | Yes |
| V-6 | F | 40’s | Asian | Primary FSGS | No | PEX, RTX | Yes |

### Primary Glomerular Cell Isolation

Human foetal kidneys were obtained from the Human Developmental Biology Resource (http://hdbr.org) at 17-21 postconceptional weeks with ethical approval from the National Health Service REC (23/LO/0312) and podocytes and glomerular endothelial cells (gEC) were isolated as previously described.^20^ Briefly, kidneys were minced and digested at 37 °C in collagenase II before passage through 100-μm sieves (3 times) and glomeruli collected on a 40-μm sieve. These were plated and grown at 37°C at 5% CO^2^ in a monolayer until confluent on 1% matrigel (Corning) in VRADD media (RPMI-1640, ThermoFisher, 218750910 supplemented with 10% FBS, 1% penicillin/streptomycin, 100 nM 1,25(OH)2D3, 1 μM all trans retinoic acid and 100 nM dexamethasone). Cells then underwent magnetic activated cell sorting (MACS) to isolate podocytes (using biotin anti-podoplanin antibody, BioLegend, 337015, conjugated to anti-biotin microbeads, Miltenyi Biotec, 130-090-485) and gECs (using anti-CD31 microbeads, Miltenyi Biotec, 130-091-935). Once sorted, podocytes were expanded in VRADD media and gECs in microvascular endothelial media MV2 (Cell Biologics, H1168). Cells were used until a maximum of passage 5.

### Glomerulus-On-A-Chip (GOAC)

We generated the GOAC in OrganoPlate 3-lane 40-well plates (Mimetas) with a modified method, based on previously described studies.^20,21^ First, a collagen matrix (5 mg/ml AMSbio Cultrex 3D Collagen-I rat tail, R&D systems 3447-020-01, 1M HEPES, 7 g/l NaHCO_3_) was generated in the middle channel of the chip before podocyte seeding 24 hours later and then gEC seeding at 72 hours (15,000 of each cell type/microchip). GOACs were cultured with MV2 in the capillary channel, and VRADD media in the urinary channel. The plate was then left for 3 days on a rocker (lateral tilt of 7° every 8 minutes to induce bidirectional flow). The cell layer was then tested for patency with the addition of 70kDa anionic flurorescein dextran (ThermoFisher D1823) to the capillary channel. One hour later, each chip was assessed for visual leak in the urinary channel using fluorescent microscopy. If patency was confirmed, each microchip was washed and then exposed to serum free endothelial media (SFEM) containing 1% healthy control or disease sera with the addition of 1% rabbit complement (Bio-Rad D2438), refreshing every 24 hours before returning to the rocker; *n* = 3 microchips/condition. At 72 hours, media in the capillary channel was replaced with SFEM containing 40 mg/ml FITC-albumin (Merck, A9771), and the urinary channel media replaced with serum-free RPMI (ThermoFisher, 218750910). One hour later, the urinary space media was collected from the inlet and outlet and FITC-albumin absorbance read at 485 nm in duplicate. Data are shown as mean ±SD for each patient across at least 2 different OrganoPlates. Replicates were standardised by calculating fold change relative to the f healthy control mean Abs_485_ run on the same plate which was given a value of 1. Fold change by the following formula:

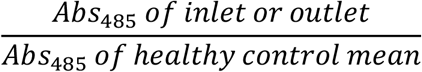

### Immunostaining

Each microchip was washed three times with 100ml PBS, then fixed with 50ml 4% PFA for 20 minutes at room temperature prior to three further washes. 50ml of CAS block (ThermoFisher 008120) was added to each well and incubated for 60 mins prior to three washes with 50ml 0.1% Triton in PBS (PBS-T). Subsequently, microchips were incubated with sheep anti-nephrin (1:100) (Biotechne AF4269) overnight at 4°C. Then, chips were washed three times with 0.1% PBS-T prior to the addition of secondary antibodies: donkey anti-sheep IgG (1:100, ThermoFisher, A-11016, Alexa Fluor 594), goat anti-human IgG (1:100, ThermoFisher, A-11013, Alexa Fluor 488) along with Hoechst (1:1000) to visualise cell nuclei for 1 hour at room temperature. Microchips were then washed three times in PBS and stored at 4°C prior to imaging.

Biopsy samples embedded in wax were rehydrated in descending concentrations of ethanol (100%-70%) before washing in Histoclear (Geneflow, HS-202). Antigen retrieval was performed using Tris-EDTA (Tris-base 1.21g/L, EDTA 0.37g/L, 0.05% Tween-20) buffer (heated for 10 minutes on high power in microwave, slides were added for a further 25 minutes on medium power) before following the antibody labelling protocol used for staining on the microchip. Biopsy slides were mounted using SlowFade mounting medium (Invitrogen, S36936). Negative controls were prepared omitting the primary antibody.

### Image Analysis

GOAC samples were imaged using an LSM710 confocal microscope (Zeiss) at 10x magnification, settings were kept consistent throughout. Biopsy samples were imaged using an Observer 7 colour light microscope (Zeiss) (x40). All images were analysed using Image J, co-localisation of IgG and nephrin was calculated by establishing a threshold for each fluorophore optimised for both positive and healthy control staining and calculating the area of overlap for each GOAC or individual glomerulus for each biopsy (mean of 7 glomeruli was calculated per biopsy). Percentage area co-localisation was calculated using the following formula:

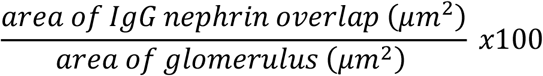

### Statistical Analysis

Statistical analyses were performed using GraphPad Prism 11.0 for Windows (GraphPad Software, Boston, MA). Data were standardised between each GOAC plate by conversion from OD (Abs 485nm) into fold change established relative to the healthy control mean OD run on each plate, which was given a value of 1. Threshold for positivity was established by running all non-recurrent and recurrent disease sera and using Youden’s index to establish a disease threshold (fold change 1.34). Those above this threshold were considered positive for a circulating factor. Data normality was assessed by Shapiro-Wilk test. Statistical differences for comparisons with more than 2 groups were analysed by 1-way analysis of variance (ANOVA) with Tukey’s multiple comparison tests for parametric data and Kruskal-Wallis test with Dunn’s multiple comparison test for non-normally distributed data. Data with 2 groups were non-normally distributed and analysed by Wilcoxon signed-rank test. Statistical significance was considered when p ≤ 0.05.

## Results

### Recurrent disease sera exposure increases GOAC permeability

To determine whether the GOAC could accurately detect the presence of a pathogenic circulating factor, it was exposed to sera from 22 participants in our inception cohort (healthy control n=4, FSGS patients with non-recurrent disease n=4, or recurrent disease n=14), with FITC-albumin leak into the urinary channel subsequently quantified, as a measure of filtration barrier integrity. We hypothesised that non-recurrent disease sera would not increase filtration barrier permeability, whereas recurrent disease sera would cause significant leak. We included a positive control with biopsy proven recurrent MCD unresponsive to B cell depletional treatment, therefore known to have a pathogenic circulating factor (Figure 1a i-iv). The positive control induced a 2.65 ± 0.09 fold increase in FITC-albumin leak compared with the healthy control, validating the assay.

**Figure 1:**
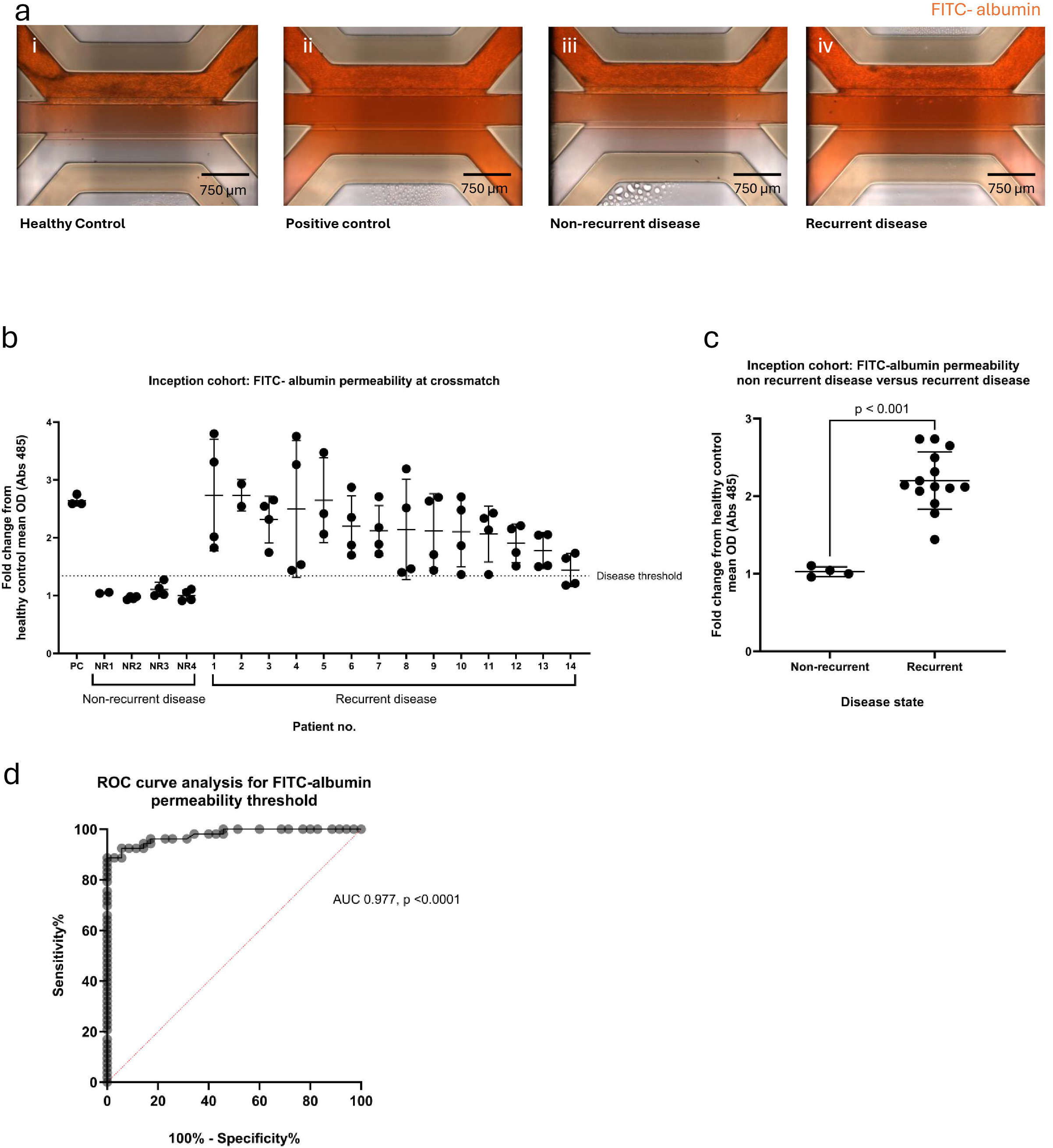
the glomerulus-on-a-chip as a diagnostic tool to predict disease recurrence. **a)** Representative images of FITC-albumin permeability in the GOAC exposed to (i) healthy control participant, (ii) positive control patient, (iii) non-recurrent disease patient, and (iv) transplantation recurrent disease patient. **b)** Quantification of inception cohort FITC-albumin permeability at crossmatch. Fold change from healthy control mean OD (Abs 485). PC – positive control, NR 1-4 – non-recurrent disease, 1-14 – recurrent disease. Disease threshold determined following ROC curve analysis and application of Youden’s index. **c)** Quantification of inception cohort mean permeability of non-recurrent disease participants versus recurrent disease participants (p<0.001). **d)** ROC curve analysis for FITC albumin permeability threshold (AUC 0.977, p<0.001). Data are shown as mean ±SD of 2-4 GOACs per patient. AUC – area under curve, FITC – Fluorescein isothiocyanate, NR – non-recurrent, PC – positive control.

First the threshold for assay positivity was established using HC sera. To do this, we used the mean and 2SD values of albumin leak to establish the upper limit of normality. Thereafter, each serum sample was compared with a HC standard run on each plate to allow results to be normalised between experiments. Albumin leak was expressed as the fold change relative to HC which was given an arbitrary value of 1. None of the 4 patients with non-recurrent disease met the HC threshold for significant FITC-albumin permeability (4/4, 100%), whereas all patients with recurrent disease surpassed the threshold (14/14, 100%) indicating the presence of a pathogenic circulating factor (Figure 1b).

Whilst the non-recurrent disease samples had a mean fold change in FITC-albumin permeability of 1.02 ± 0.06, it was significantly higher in the recurrent disease samples with a mean fold change of 2.20 ± 0.37 (*p<*0.001) (Figure 1c). ROC curve analysis of FITC-albumin permeability data between non-recurrent and recurrent disease cohorts confirmed strongly discriminatory ability of the GOAC to predict recurrence, with an area under the curve (AUC) of 0.98 (P < 0.001) (Figure 1d). Using the ROC analysis and Youden’s index allowed us to determine the optimal diagnostic threshold of 1.34 (sensitivity 89% (CI 77.4-94.7%), specificity 100% (CI 91.1-100%), which we implemented thereafter for the validation cohort.

When comparing the outcomes between men (n=9) and women (n=5) we found a significantly increased leak in men compared with women (mean fold increase 2.16 ± 0.07 compared to 1.83 ± 0.08 respectively, p= 0.03). It is not clear what would explain this difference as no sex bias has been previously reported for post-transplant FSGS recurrence and there were no significant differences in demographic features associated with greater risk of relapse, such as age or white ethnicity.

### IgG-nephrin co-localisation indicates the presence of anti-nephrin antibodies

To establish whether the pathogenic circulating factor was antibody-associated or otherwise we proceeded to immunostaining, choosing to focus on co-localisation of IgG positive signal, and nephrin positive signal as the most widely reported slit diaphragm antigen target (Figure 2a i-iv). First, we established a threshold of 1.148% for co-localisation using serum from remission patients (mean percentage co-localisation found in patients I2 and I12). In 9/13 recurrent DP patients (69%), IgG and nephrin co-localised to levels above our threshold indicating the presence of anti-nephrin antibodies. Importantly, 4/13 (30.7%) had co-localisation percentages lower than the threshold, indicating a potential non-nephrin antibody-associated process (Figure 2b). Percentage co-localisation was consistent with individual clinical response to B cell-targeting therapy, those with co-localisation above threshold, experienced at least partial response to treatment, whereas those below threshold had been refractory to treatment returning to dialysis.

**Figure 2:**
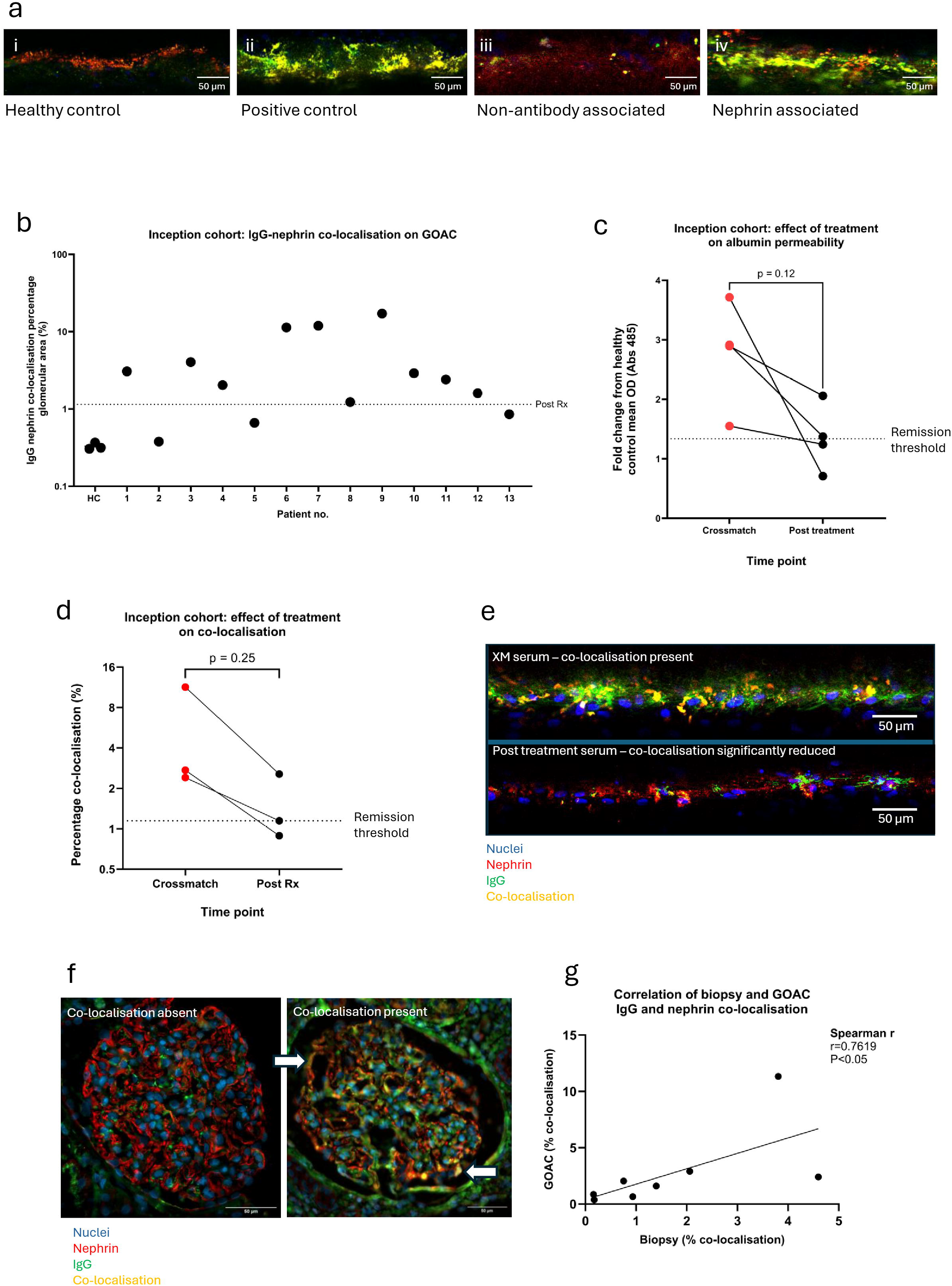
the glomerulus on a chip identifies anti-nephrin antibodies in recurrent disease. **(a)** Representative images of GOAC exposed to sera from (i) healthy control participant, (ii) positive control patient, (iii) non-nephrin antibody associated recurrent disease patient, and (iv) nephrin antibody associated recurrent disease patient. Nephrin and human IgG are visualised in red and green respectively, and co-localisation is shown in yellow. Quantification of **(b)** the inception cohort nephrin-IgG co-localisation on the GOAC, **(c)** effect of treatment on FITC-albumin permeability and **(d)** nephrin-IgG co-localisation. Representative images of **(e)** GOAC nephrin-IgG co-localisation pre and post treatment of the same patient, and **(f)** glomerular biopsies from two patients with and without nephrin-IgG co-localisation. **(g)** Spearman’s correlation analysis of GOAC versus biopsy nephrin-IgG overlap from patients where both biopsy and GOAC images were available. GOAC – glomerulus on a chip, Rx – treatment, XM – crossmatch.

### B cell targeted therapy results in reduction in GOAC albumin permeability and IgG deposition

We obtained additional post-treatment sera from four inception cohort recurrent disease patients and compared albumin permeability and IgG deposition pre- and post-treatment. At the time of biopsy proven recurrence, all patients received five sessions of PEX and 2g Rituximab. All post treatment sera demonstrated a reduction in fold change albumin permeability (Wilcoxon p=0.12) (Figure 2c). Albumin permeability for three participants dropped below our threshold corresponding with complete clinical remission. These three patients were found to have significant IgG nephrin co-localisation, which for two reduced to below threshold following treatment (Wilcoxon, p=0.25) (Figure 2d,e). One participant remained above threshold for albumin permeability fold change and continued to experience an active disease state. However, this individual showed no co-localisation of IgG and nephrin, suggesting they may represent patients with a non-nephrin antibody mediated circulating factor.

### In vitro IgG-nephrin co-localisation is consistent with transplant biopsy co-localisation

We compared our GOAC immunostaining with biopsy samples available for 8/13 of the recurrent DP patients (Figure 2f). There was a positive correlation between GOAC and *in vivo* percentage co-localisation (R=0.76, p<0.05) (Figure 2g) suggesting the GOAC could be a reliable substitute in circumstances where a biopsy was not available.

### GOAC pipeline accurately predicted clinical outcomes in validation cohort

Finally, we tested an external validation cohort of six patients blinded for clinical outcome in our GOAC pipeline. We initially tested for increased FITC-albumin permeability as an indicator of the presence of a damaging circulating factor. All 6 patient sera induced albumin leak into the urinary channel above the defined threshold with a mean fold change of 2.216 ± 0.27, strongly indicative of damaging circulating factors in the serum milieu (Figure 3a). Additionally, we found co-localisation of IgG and nephrin in 3/6 of patients indicating that they should respond well to B cell-targeting therapy. Notably one patient was on the threshold borderline (Figure 3b).

**Figure 3:**
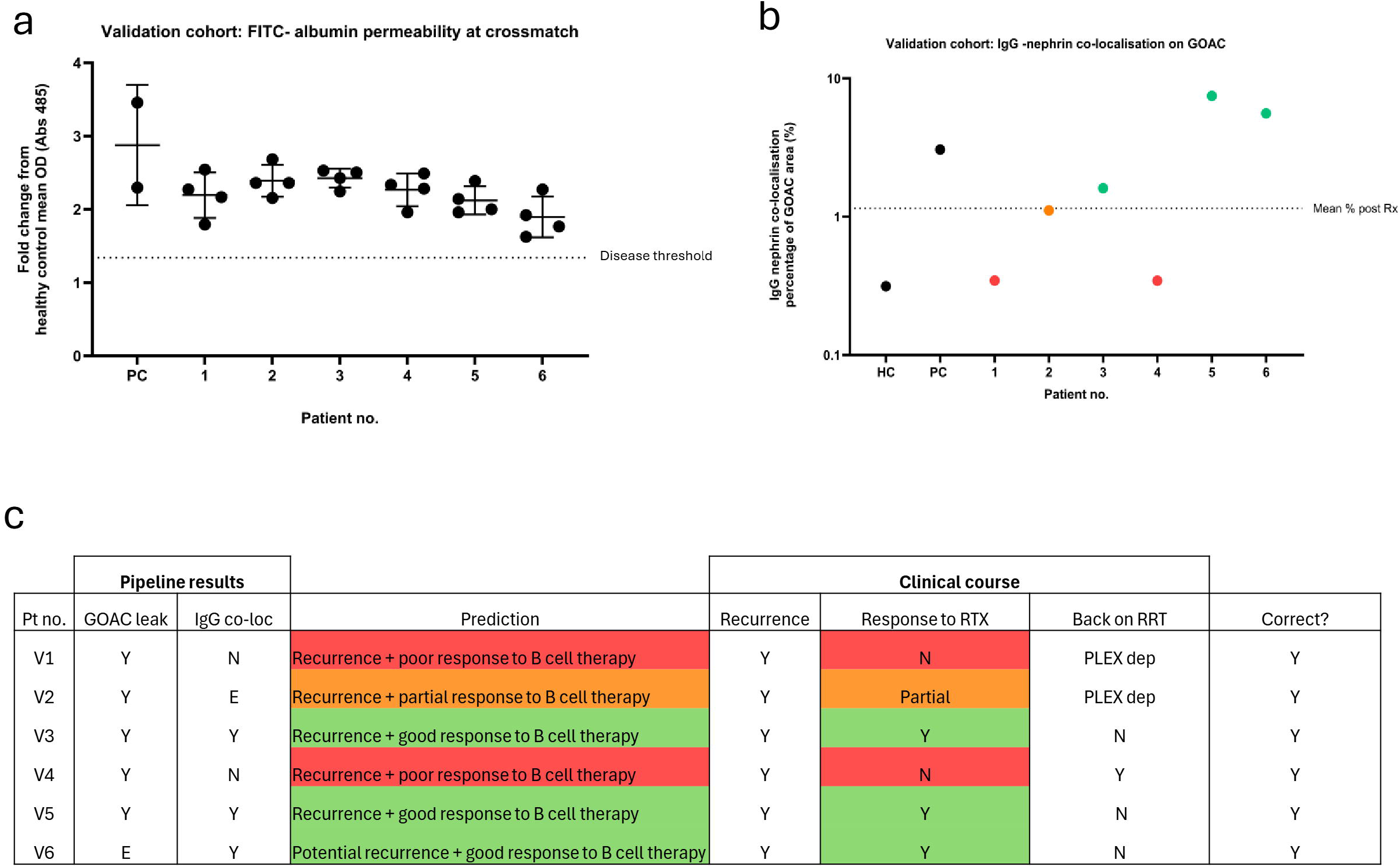
the glomerulus-on-a-chip accurately predicts disease recurrence and response to treatment in a blinded cohort. Quantification of (a) FITC-albumin fold change and (b) IgG-nephrin co-localisation in patients whom we were blinded to clinical characteristics. (c) Our post-experimental predictions prior to unblinding clinical characteristics, with the outcome of our predictions (6/6). GOAC – glomerulus on a chip, RRT – renal replacement therapy, RTX – rituximab.

We then unblinded the clinical outcomes. Strikingly, all 6 patients experienced disease recurrence, reflected in our FITC-albumin assay. The three patients who were above the threshold for IgG-nephrin co-localisation had responded well to B cell therapy, and the patient who was on the threshold line, displayed a partial response to B cell therapy with stabilised function but PEX dependency. The other 2 patients showed no clinical response to RTX and showed no IgG/nephrin co-localisation in our GOAC, one remains PEX dependent and one has now returned to dialysis. The blinded cohort *in vitro* outcomes matched (6/6) with clinical outcomes (Figure 3c), providing strong validation for our diagnostic pipeline.

Collectively, we provide a novel diagnostic pipeline for patients with glomerular disease based on current state-of-the-art *in vitro* technology. This pipeline could stratify treatment regime, lowering the risk of recurrence in patients, and reducing graft rejection (Figure 4).

**Figure 4.**
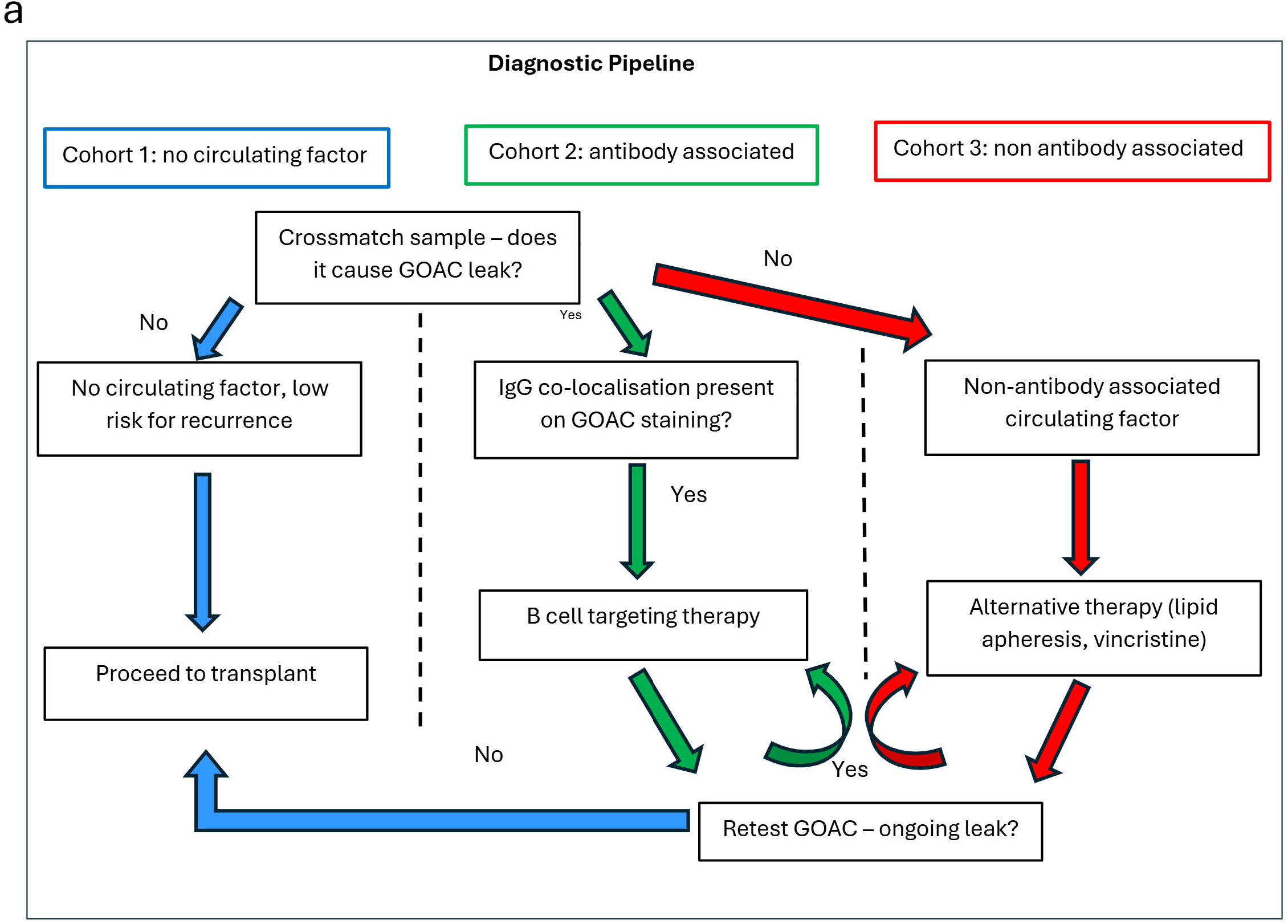
Clinical diagnostic pipeline using the GOAC. a) A diagnostic pipeline to aid therapeutic stratification of patients as risk of recurrent diffuse podocytopathies prior to transplantation.

## Discussion

This pipeline has the potential to transform our approach towards recurrent DPs by employing a modified GOAC model to screen for a pathogenic circulating factor, subsequently defining it as a nephrin antibody-associated process or otherwise, to inform therapeutic strategy surrounding transplantation.

To our knowledge, this work is the first to use recurrent podocytopathy sera in the GOAC model. We have demonstrated excellent sensitivity and specificity of our pipeline in determining the presence of a pathogenic circulating factor both at the point of crossmatch and post treatment. Previous work has focused on defining damage to the recapitulated filtration barrier by exposing it to sera from native glomerular disease. In these studies, membranous nephropathy, IgA nephropathy and lupus nephritis demonstrated increased permeability and loss of podocyte specific cell markers.^22–24^ However, none have shown consistent disease effect with FSGS sera apart from Mason et al.^22–24^ In our study, we have modified the methodology by introducing a patency confirming step and added complement to enhance podocyte injury in the presence of antibody. These may explain the enhanced reliability of our results.

Our prevalence of antibody-associated disease and response to treatment in recurrent DPs is consistent with published international data in adults.^1^ Reliable methodology which is reproducible across laboratories to detect serum anti-slit diaphragm antibodies both pre- and post-transplantation remains elusive. Immunoprecipitation-western blot is currently the most widely-used technique but is both labour intensive and only semi-quantitative.^9,12,25^ Immunoprecipitation-ELISA (IP-ELISA) has also been implemented with the aim of streamlining identification and quantifying antibody titres.^4,13,26,27^ However, specificity of results is dependent on chosen nephrin derivative, with human recombinant nephrin associated with the lowest rate of false positive results and murine-derived the highest, likely secondary to cross reactivity with anti-alpha galactase (anti-alpha-GAL) antibodies.^9,10,28^ Pecoraro et al. investigated this further reporting no consistent anti-nephrin antibody disease enrichment using IP-ELISA despite eliminating anti-alpha-GAL antibodies.^10^

A recent publication used biopsy immunostaining in conjunction with serological testing to report prevalence of anti-slit diaphragm antibodies in a mixed adult/paediatric recurrent FSGS cohort.^5^ They reported presence of anti-podocin and anti-KIRREL-1 antibodies, some in conjunction with anti-nephrin antibodies, but none were found to be positive for anti-nephrin antibodies alone ^5^, suggesting perhaps focus in the field should shift towards testing for these in our patient sera. In this regard, our methodology could easily be adapted to screen for other immunoglobulin subtypes.

Current literature is predominately descriptive reporting presence or absence of antibodies in disease or remission states, but this does not necessarily confirm pathogenicity. In addition, there is reported correlation between the presence of anti-nephrin antibodies and favourable response to steroid and B cell directed therapy.^8,11,29–32^ Whilst this is useful in helping to inform likelihood of clinical trajectory, it is not patient specific which is key to guiding appropriate treatment in a clinically heterogenous condition. Our pipeline is an advancement on this existing work; confirming pathogenicity through destruction of the recapitulated filtration barrier; defining the nature of the factor for every patient; and confirming response to treatment. Perhaps, most importantly, we used the GOAC with high predictive accuracy with both disease recurrence and response to treatment in a validation cohort. This predictive element adds value to our model from a functional tool to confirm disease state to one that facilitates pre-emptive clinical decision making.

Given the predictive strengths of our system, we believe that it could be implemented in the pre-transplant assessment setting. In this context, it could be used to detect a circulating pathogenic factor and confirm its removal before transplant waitlisting, during the waiting period or prior to living-donor transplantation, minimising the risk of post-transplant recurrence (Figure 4). This strategy has been positively received by patient groups, during our public and patient engagement work, who feel reassured that transplantation decisions would occur alongside a biomarker assessment suggesting a reduced risk of disease recurrence.

Additionally, our modified GOAC platform could be used to test drug repurposing and novel therapeutic options in recurrent podocytopathy patients with non-antibody associated disease, as shown in our previous work with vincristine.^20^ We studied sera from a patient with a native FSGS, who had been clinically unresponsive to B cell directed therapy and whose sera caused increased GOAC permeability despite IgG depletion. We subsequently tested sera post-vincristine therapy, demonstrating reduction in albumin permeability as well as podocyte cytoskeleton remodelling correlating with clinical remission. In addition, vincristine treatment of sera from another patient with native FSGS proved ineffective in reducing GOAC leak, showing the potential for the pipeline to protect patients from receiving ineffective treatments with significant side effect profiles. Confirming treatment efficacy of alternative therapies *in vitro* would be invaluable and more detailed exploration of podocyte remodelling may provide further insight into non-antibody associated disease mechanism in individual patients, laying groundwork for future research.

Lastly, the ability of our model to replicate human tissue architecture *in vitro* and predict antibody associated disease has the potential to replace the need for a biopsy, which may prove particularly valuable in high-risk post-operative settings.

This is an exploratory study, limited by sample size. The use of primary cells, whilst chosen for their close representation of the *in vivo* glomerular environment, adds a degree of variability which is possibly reflected in the variance of our data. However, we have minimised this as much as possible through normalising responses on each plate to those of healthy controls.

In conclusion, our work presents a novel and pragmatic stratification pipeline which is directly applicable to clinical practice. It can inform pre-emptive and post-recurrence therapeutic strategy as well as providing confirmation of disease remission. This precision medicine approach aims to revolutionise our management of those at risk of recurrent podocytopathies, optimising their graft function and survival.

## Disclosure statement

the authors have declared that no conflict of interest exists.

## Data sharing statement

all data supporting the findings of this study are included within the article and its supplementary materials.

## Data Availability

All data produced in the present study are available upon reasonable request to the authors

## Acknowledgments

we are grateful to all the patients who participated in the study. We would like to thank Dr Dale Moulding of the UCL Great Ormond Street Institute of Child Health for assistance and expertise with microscopic imaging.

## Funding support

Kidney Research UK, Stoneygate Michael Nicholson Award (002-20241015), Wellcome Trust Investigator Award (220895/Z/20/Z to DAL), NIHR Biomedical Research Centre at Great Ormond Street Hospital for Children NHS Foundation Trust and University College London through a Career Development Catalyst Fellowship Award to WJM, LifeArc-Kidney Research UK Centre for Rare Kidney Diseases and The Human Developmental Biology Resource funded by the Medical Research Council and the Wellcome Trust.

## Notes

### Competing Interest Statement

The authors have declared no competing interest.

### Author Declarations

Human foetal kidneys were obtained from the Human Developmental Biology Resource with ethical approval from the National Health Service REC (23/LO/0312) All patient samples were obtained with ethical approval (05/Q0508/6 Royal Free Hospital Research Ethics Committee)

## References

1. Uffing A, Pérez-Sáez MJ, La Manna G, et al. A large, international study on post-transplant glomerular diseases: the TANGO project. BMC Nephrol. 2018;19(1):229. doi:10.1186/s12882-018-1025-z

2. Uffing A, Pérez-Sáez MJ, Mazzali M, et al. Recurrence of FSGS after Kidney Transplantation in Adults. Clinical Journal of the American Society of Nephrology. 2020;15(2):247–256. doi:10.2215/CJN.08970719

3. Hayward S, Parmesar K, Saleem MA. What is circulating factor disease and how is it currently explained? Pediatr Nephrol. 2023;38(11):3513–3518. doi:10.1007/s00467-023-05928-8

4. Hengel FE, Dehde S, Lassé M, et al. Autoantibodies Targeting Nephrin in Podocytopathies. New England Journal of Medicine. 2024;391(5):422–433. doi:10.1056/NEJMoa2314471

5. Raglianti V, Angelotti ML, Cirillo L, et al. Anti-podocin and Anti-KIRREL1 Antibodies and Steroid Resistance, FSGS, and Disease Recurrence after Transplantation in Autoimmune Podocytopathies. J Am Soc Nephrol. Published online July 23, 2026. doi:10.1681/ASN.0000001195

6. Raglianti V, Angelotti ML, De Chiara L, et al. Anti-slit Antibodies against Podocin and Kirrel1 in Pediatric and Adult Podocytopathies. Journal of the American Society of Nephrology. 2025;36(4):702–705. doi:10.1681/ASN.0000000642

7. Watts AJB, Keller KH, Lerner G, et al. Discovery of Autoantibodies Targeting Nephrin in Minimal Change Disease Supports a Novel Autoimmune Etiology. Journal of the American Society of Nephrology. 2022;33(1):238–252. doi:10.1681/ASN.2021060794

8. Hengel FE, Dehde S, Yilmaz A, et al. Anti-nephrin autoantibodies in steroid-resistant nephrotic syndrome may inform treatment strategy. Kidney Int. 2025;107(6):1099–1103. doi:10.1016/j.kint.2025.01.019

9. Liu P, Liu S, Dalal V, et al. Evaluation of Methodologies in Anti-nephrin Autoantibody Detection. bioRxiv. Published online July 25, 2024. doi:10.1101/2024.07.25.605154

10. Pecoraro F, Perico L, Casiraghi F, et al. Anti-nephrin antibodies are not enriched in patients with primary and post-transplant recurrent podocytopathies. Journal of Clinical Investigation. Published online April 28, 2026. doi:10.1172/JCI204727

11. Raglianti V, Angelotti ML, De Chiara L, Romagnani P. Anti-nephrin, anti-podocin and anti-Kirrel1 antibodies: Biological challenges and clinical implications. Nephrology Dialysis Transplantation. Published online August 15, 2025. doi:10.1093/ndt/gfaf156

12. Hattori M, Shirai Y, Kanda S, et al. Circulating nephrin autoantibodies and posttransplant recurrence of primary focal segmental glomerulosclerosis. Am J Transplant. 2022;22(10):2478–2480. doi:10.1111/ajt.17077

13. Weins A, Hamroun A, Provot F, Keller KH, Gibier JB, Watts AJ. Pre-Transplant Nephrin Autoantibodies Predict Post-Transplant Recurrent Focal Segmental Glomerulosclerosis (FSGS). Journal of the American Society of Nephrology. 2023;34(11S):309–309. doi:10.1681/ASN.20233411S1309d

14. Lafargue MC, Hullekes F, Leon J, et al. Anti-Nephrin Antibodies in Focal Segmental Glomerulosclerosis Recurrence After Transplantation: A Multicenter Study from the TANGO Consortium. American Journal of Transplantation. Published online November 2026. doi:10.1016/j.ajt.2026.11.007

15. Angeletti A, Bin S, Magnasco A, Bruschi M, Cravedi P, Ghiggeri GM. Efficacy of combined rituximab and daratumumab treatment in posttransplant recurrent focal segmental glomerulosclerosis. American Journal of Transplantation. 2024;24(4):688–692. doi:10.1016/j.ajt.2023.12.010

16. Avaniadi PD, Akifova A, Schrezenmeier EV, et al. Daratumumab as rescue therapy in refractory recurrent FSGS after kidney transplantation: a case series. Transplant International. 2026;39. doi:10.3389/ti.2026.16693

17. Leon J, Ducrocq E, Lafargue MC, et al. Anti-CD38/Anti-CD20 Rescue Therapy for Posttransplant Recurrent FSGS. Kidney Int Rep. 2026;11(9):106668. doi:10.1016/j.ekir.2026.106668

18. Aleš Rigler A, Belčič Mikič T, Bošnjak M, Arnol M. Subcutaneous Daratumumab in Recurrent Posttransplant Focal Segmental Glomerulosclerosis: A Report of 2 Cases. Kidney Med. 2026;8(9):101473. doi:10.1016/j.xkme.2026.101473

19. Randone P, Sanna E, Dolla C, et al. Rescue with obinutuzumab and daratumumab as combined B cell/plasma cell targeting approach in severe posttransplant focal segmental glomerulosclerosis recurrence. American Journal of Transplantation. 2024;24(10):1896–1900. doi:10.1016/j.ajt.2024.06.010

20. Mason WJ, Chandler JC, Gage AM, et al. Vincristine Treatment Protects Against Podocyte Damage in Focal Segmental Glomerulosclerosis. Kidney Int Rep. 2025;10(11):3998–4011. doi:10.1016/j.ekir.2025.08.002

21. Petrosyan A, Cravedi P, Villani V, et al. A glomerulus-on-a-chip to recapitulate the human glomerular filtration barrier. Nat Commun. 2019;10(1):3656. doi:10.1038/s41467-019-11577-z

22. Perin L, Da Sacco S. Generation of a Glomerular Filtration Barrier on a Glomerulus-on-a-Chip Platform. Methods Mol Biol. 2022;2373:121–131. doi:10.1007/978-1-0716-1693-2_8

23. Kim SY, Choi YY, Kwon EJ, et al. Characterizing Glomerular Barrier Dysfunction with Patient-Derived Serum in Glomerulus-on-a-Chip Models: Unveiling New Insights into Glomerulonephritis. Int J Mol Sci. 2024;25(10):5121. doi:10.3390/ijms25105121

24. Doi K, Kimura H, Matsunaga YT, Fujii T, Nangaku M. Glomerulus-on-a-Chip: Current Insights and Future Potential Towards Recapitulating Selectively Permeable Filtration Systems. Int J Nephrol Renovasc Dis. 2022;15:85–101. doi:10.2147/IJNRD.S344725

25. Vivarelli M, Colucci M. Anti-nephrin antibodies in recurrence of focal segmental glomerulosclerosis: closer to discovering the Holy Grail? Kidney Int. 2024;105(3):440–442. doi:10.1016/j.kint.2023.12.015

26. Watts AJB, Keller KH, Lerner G, et al. Discovery of Autoantibodies Targeting Nephrin in Minimal Change Disease Supports a Novel Autoimmune Etiology. Journal of the American Society of Nephrology. 2022;33(1):238–252. doi:10.1681/ASN.2021060794

27. Shirai Y, Miura K, Ishizuka K, et al. A multi-institutional study found a possible role of anti-nephrin antibodies in post-transplant focal segmental glomerulosclerosis recurrence. Kidney Int. 2024;105(3):608–617. doi:10.1016/j.kint.2023.11.022

28. Shirai Y, Miura K, Morimoto K, Yokoo T, Hattori M. Reevaluating anti-nephrin autoantibodies by ELISA using human embryonic kidney–derived recombinant extracellular domain of human nephrin. Kidney Int. 2025;107(4):757–759. doi:10.1016/j.kint.2025.01.003

29. Fernandez H, Canetta P, Bomback A, et al. Monitoring anti-nephrin antibodies in the management of recurrent diffuse podocytopathy. Kidney Int. Published online June 2026. doi:10.1016/j.kint.2026.06.013

30. Horinouchi T, Nagano C, Watts AJB. Anti-nephrin antibodies in steroid-sensitive nephrotic syndrome in Japanese children. Pediatr Nephrol. 2024;39(1):337. doi:10.1007/s00467-023-06107-5

31. Liaukouskaya N, Hengel FE, Mühlig A, Tomas NM, Huber TB. A New Hope for Treating Podocytopathies - Emerging Role of Anti-Nephrin Antibody. Clin J Am Soc Nephrol. Published online January 21, 2026. doi:10.2215/CJN.0000001008

32. Hengel FE, Beldi-Ferchiou A, Sakhi H, et al. Effect of Steroids on Anti-nephrin Autoantibodies and B cells in Minimal Change Disease. Kidney Int Rep. 2026;11(4):103794. doi:10.1016/j.ekir.2026.103794

